# Simulation model for productivity, risk and GDP impact forecasting of the COVID-19 portfolio vaccines

**DOI:** 10.1101/2020.11.01.20214122

**Authors:** Vladimir Shnaydman

## Abstract

The paper presents the methodology and modeling results for COVID-19 vaccines portfolio forecasting, including R&D output (rate and likelihood of approvals at a vaccine technology platform level) and manufacturing production output to meet worldwide demand.

In order to minimize the time and risk of global vaccination, scaling up of Operation Warp Speed (OWS) and other programs could be very beneficial, leading to increased financing for additional vaccine development programs, in both Phase III clinical trials and in manufacturing. It would also lead to a reduction of the global production time for world vaccination, from 75 months for a baseline scenario to 36 months, reducing potential global GDP loss by as much as US$4.2 trillion (US ∼ $1 trillion) when compared to the baseline scenario.

## Introduction

COVID-19 vaccine developers are racing against time to develop, test, approve and produce an effective vaccine. Currently more than 200 vaccines are under development^1^ versus the 80 to 100 programs that were underway in April 2020. Many programs are still in the preclinical stage, but more than 40 candidates are in clinical trials^2^. It is expected that several vaccines may be approved within months. Each vaccine development program belongs to one of eight technology platforms. Vaccine manufacturers have announced cumulative manufacturing capacity that could produce as many as one billion doses by the end of 2020 and up to 9 billion doses by the end of 2021 [3].

Instead of managing COVID-19 vaccine development programs individually, it was proposed in April 2020 [4], and in May 2020 [5] that portfolio concepts and methodologies [6] can be applied to the programs. This approach would lead to more coordinated planning and management of development projects, and a more efficient way to achieve strategic goals.

The paper will describe portfolio simulation as an effective tool to forecast achievability of COVID-19 portfolio strategic goals, including risk mitigation to boost vaccines development and production.

Portfolio strategic objectives are:

- The manufacture of 2 billion doses by the end of 2021, earmarked for health system workers and for people over 65.
- The manufacture of 11.2 billion doses in the shortest time with minimal risk (11.2 billion doses would vaccinate 70 per cent of the world population of 8 billion, at two doses per person) [7]. The authors [8] assume production of 18 billion doses, taking into consideration 20 per cent loss; global distribution companies are planning for 20 billion doses [9]^1^.
- Other intermediate goals could be established as well.

### Overview

In March and April 2020 it became obvious that COVID-19 vaccine development programs became numerous enough to be analyzed as a portfolio [4]. The model, published in April 2020, presented rather conservative estimates for the portfolio risk. At that time, “there is a ∼40% chance that no vaccine is approved within 18 months, a ∼67% chance that no more than one vaccine is approved, and a ∼93% chance that no more than two vaccines are approved.” [4] Then the Center for Global Development (CGD) followed, and published several blogs on the topic between June and September [10-12], and a report [8], describing a very similar methodology.

Since April 2020, the portfolio of COVID-18 vaccines moved fast [13]. Hence, the R&D portfolio risk profile is more favorable than in the model published in April [4]. At the same time, new questions emerged related to the model’s granularity, manufacturing of vaccines, and risk mitigation strategies.

### Methodology and the model

The COVID-19 vaccine supply chain includes three major blocks (Figure 1): (1) R&D and clinical trials; (2) manufacture of the vaccines, and (3) distribution of the vaccines. Each block has multiple components or sub-blocks. This paper covers only two sub-models: R&D, and clinical research and manufacturing) due to absence of data about vaccines distribution.

**Figure 1.**
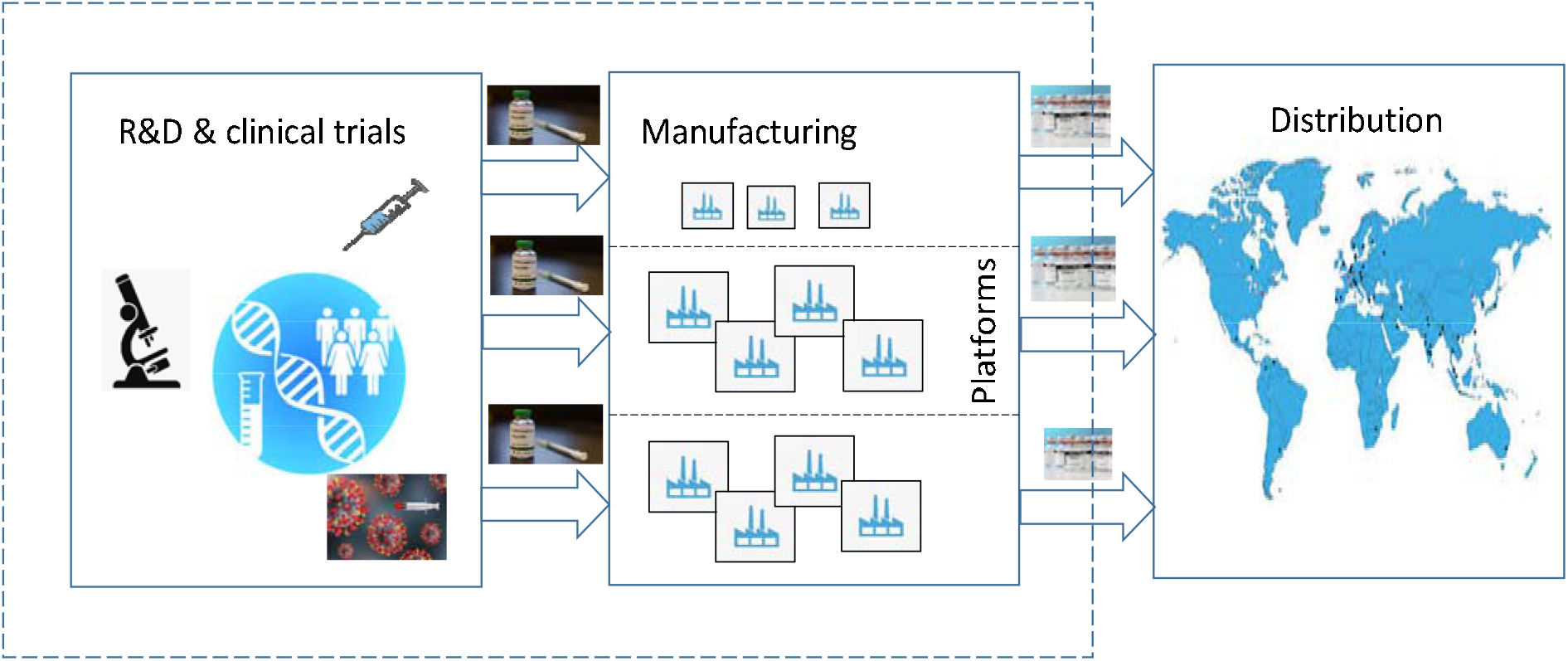
COVID-19 supply chain (Shnaydman, 2020).

The R&D/clinical research sub-model simulates a vaccine development workflow from preclinical to approval. The simulator utilizes [3] decision tree diagram with binary stochastic outcome (success or failure) [13]. The model [13] was modified to simulate the portfolio of COVID-19 vaccines. Comparing to [4], the current version of the model presents a more granular view of the vaccine R&D development process at a technology platform level and simulates production of the vaccines. It also incorporates algorithms for simulating interdependence between vaccines and speeding up Phase III trials across the portfolio to mitigate production risk and reduce production time to reach vaccination goals. Modeling capabilities are limited by data availability and data accuracy across the portfolio.

### Input data^2^

Input data for the model was collected from a variety of sources, including databases, academic and trade publications, webinars, podcasts, private communications, and others. Some data reflect a subjective view of the researchers. Therefore, the requirements for data granularity should be balanced with its uniformity across portfolio and sub-models.

#### Major data blocks

1. Portfolio landscape – number of preclinical, Phase I, II, and III candidates (approximately mid-July - August 2020) across eight platforms [1].
2. Planning horizon is 60 months. Modeling interval is a month.
3. Platform specific data for each vaccine
  a. Cycle time for each clinical phase and each vaccine
  b. Transitional POS at a phase and a platform level
  c. Integrated monthly manufacturing productivity for each platform
  d. Vaccine demand for each goal
  a. *Cycle times* were significantly compressed due to development urgency. For lead candidates [15, 16], clinical trial stages are even more compressed and overlapped because of availability of additional funding in order to maximize the speed of development. The result is that much development work is done in parallel and at risk.
  b. Probabilities of success (POS) POS is one of most important drivers of vaccine portfolio productivity. The first release of the model [4] did not differentiate phased POS across vaccine platforms due to absence of data. This assumption was in line with POS data published in several sources at that time, such as [17-21]. In these papers and reports, integrated POS for all vaccines [18, 19] and vaccines for infectious diseases [20] were calculated based on statistical processing of historical data obtained from previous clinical trials. However, the lack of data needed to calculate POS at a platform level for COVID-19 vaccines portfolio required another approach.

#### Elicitation of POS for vaccine candidates across technology platforms

The three major approaches used to derive POS are summarized in Table 1:

**Table 1.**
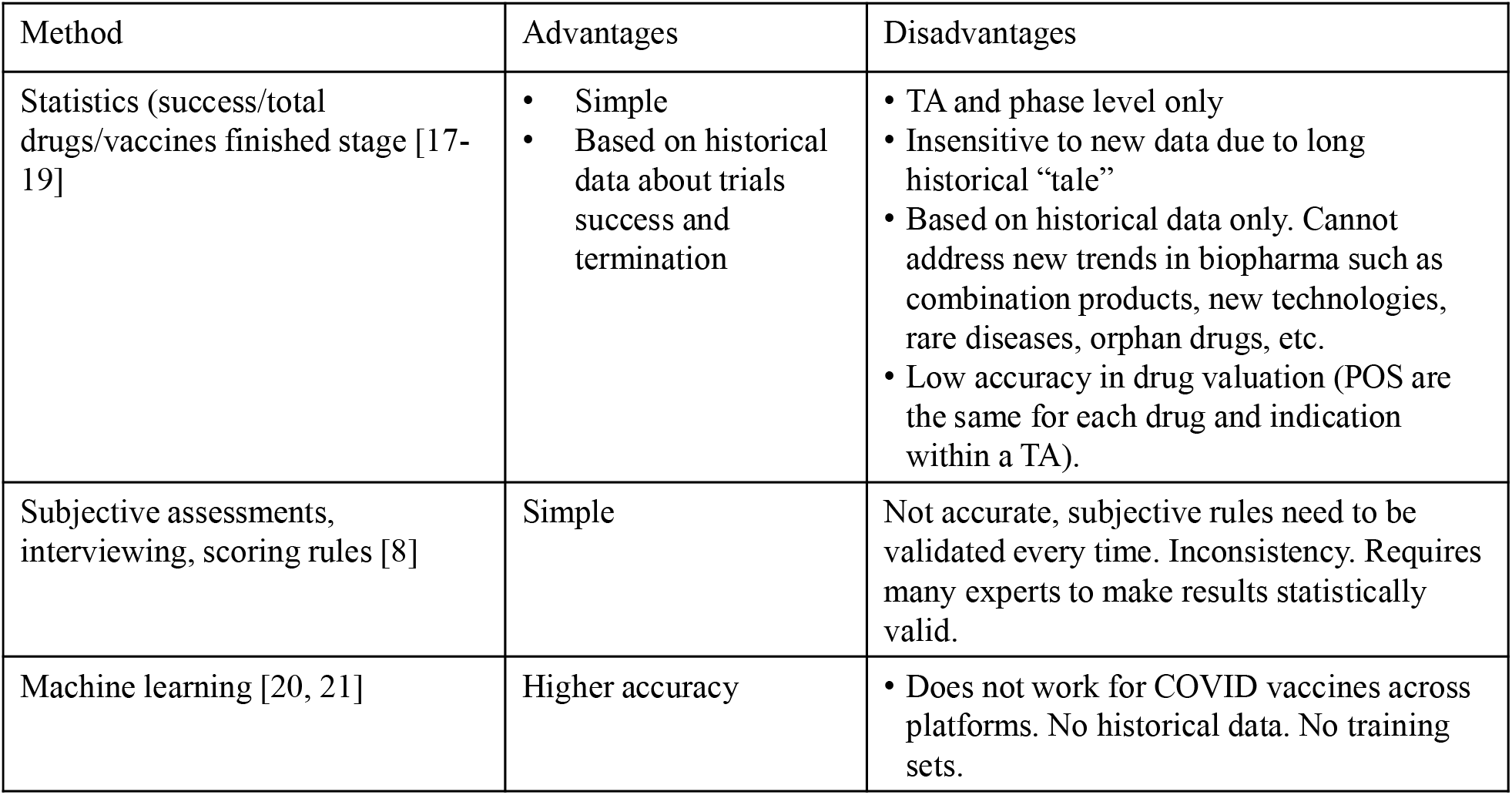
Advantages and disadvantages of different techniques for POS elicitation.

For accurate simulation of COVID-19 vaccines portfolio, POS elicitation needs to be derived for each platform and each phase of a clinical trial. As the analysis in Table 1 indicates, currently available techniques may not be effective. For example, experts interviewing requires a large number of experts (16 in [8]) in order to make results statistically significant. Also, COVID-19 vaccines development portfolio is changing so rapidly that making another round of relevant interviews with the same large team of experts could be problematic.

Therefore, a new approach was proposed due to lack of historical data: POS elicitation from the development risk prospective. The approach is based on the assumption that the higher the POS, the lower development and regulatory risk, and vice versa. In other words, how can one convert development and regulatory risk into POS?

It looks very beneficial to evaluate development risk based on qualitative pairwise comparison of different risks related to global categories such as safety and efficacy at a platform level. Then, platforms are ranked according to risks related to each category, and POS derived according to ranked platforms using Analytic Hierarchy Process (AHP) - a multiple criteria decision making tool that has been widely used by in many applications and verticals [22-24], such as planning, resource allocation, and risk management.

An example of structured risk analysis for COVID-19 vaccine development using AHP is presented in Figure 2.

**Figure 2.**
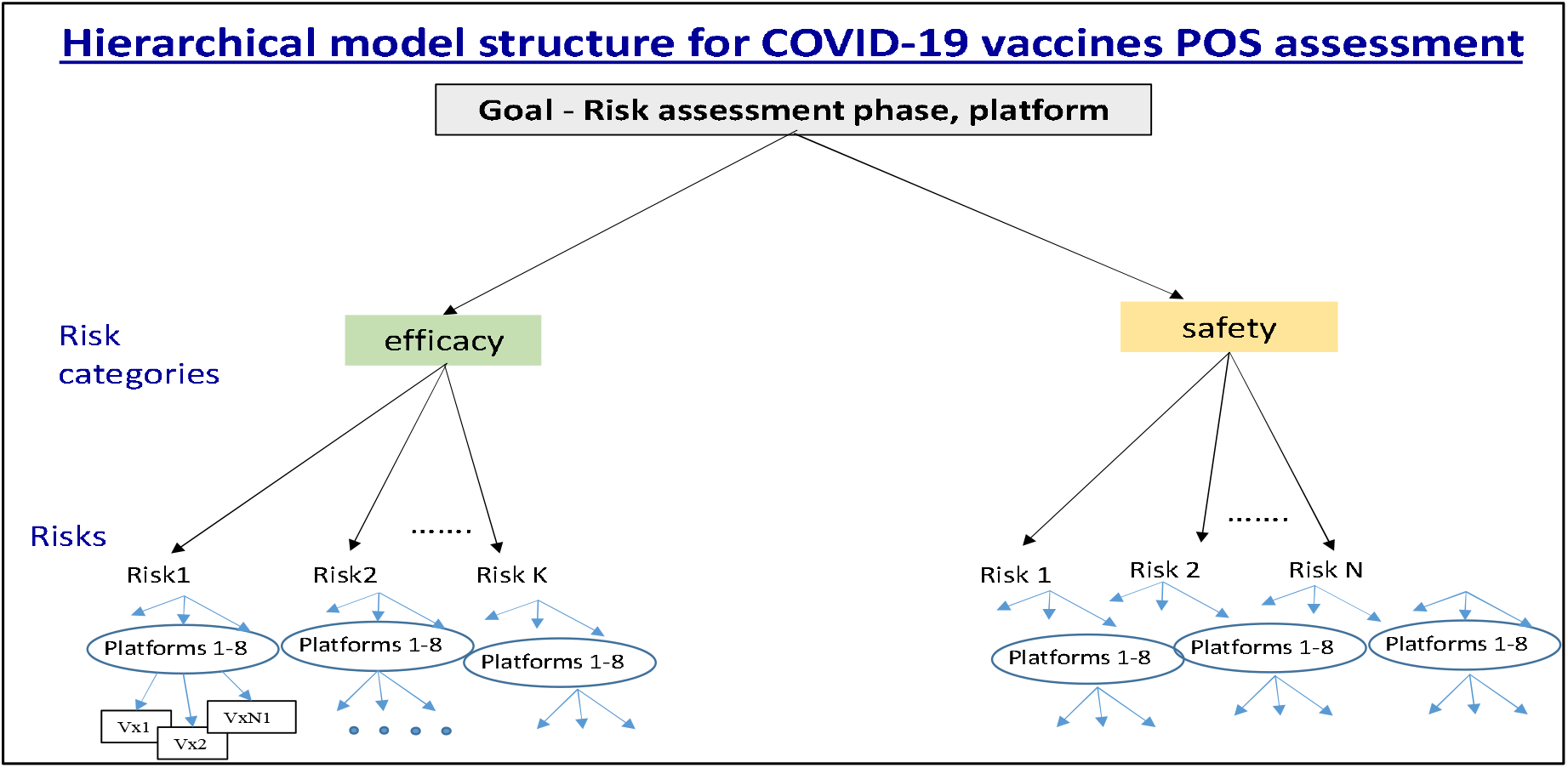
Risk hierarchical structure for POS calculation in vaccine development (Shnaydman, 2020)

The highest level of hierarchy, “Goal - Risk assessment across technology platforms and phases” is comprised of *risk categories*, such as efficacy and safety. Each category includes several most significant risks. Risk assessment includes two steps. The first step is a pairwise comparison of *risk categories* at a lexicographic scale from one to nine by an expert or a group of experts to determine weight of each category according to the goal. Pairwise comparison of risk categories generates their weights. For example, a vaccine efficacy of 0.4 and safety of 0.6; the sum should be equal to 1. Then, at the second step, pairwise comparison of risks within each risk category for each platform is conducted.

AHP software calculates ranking of each vaccine platform. In addition, experts estimated the minimum and maximum POS for each platform and phase, and the POS for each platform and phase was calculated^3^.

Risk assessment in vaccine development is subjective. In order to avoid bias, a group of experts is preferred to make robust assessments [22-24]. Also, AHP software is able to report inconsistencies in experts’ judgements which interviewing lacks. Sensitivity to POS variations for the COVID portfolio simulator was tested in [4].

### Manufacturing productivity and vaccine demand

Several manufacturers have announced their plans to produce about one billion COVID-19 vaccine doses by the end of 2020 and about eight to nine billion doses by the end of 2021 [3, 25]. Not all vaccines are expected to be successful, so production volumes should be adjusted for POS.

The model incorporates an integrated network of manufacturing facilities for each platform, and is characterized by its productivity. For example, 25 facilities participate in manufacturing of COVID-19 vaccines [26-28].

4. Algorithms describing interdependence between vaccines.

Each platform includes multiple vaccine development programs. Vaccines within a platform have a lot of similarities, but they are still different. Therefore, the risk profile of each vaccine is unique, but the failure or success of a vaccine may have an impact on other candidates within a platform. *Vaccine success* may reduce risk for other vaccines within a platform and, therefore, increase their POS. *Vaccine failure* will increase risk for other vaccines, and therefore will reduce their POS.

It is assumed in the model that after each successful clinical trial, POS for other trials within a phase increases by *x* %, and decreases by *y* % if a clinical trial fails. This algorithm aligns with current practices of portfolio analysis for interdependent drugs, summarized in [14, 17]. Parameters *x* % and *y* % are derived based on an expert knowledge^4^. POS may be increased up to maximum value of POS (POS_max_) for a platform/phase, and will be decreased to the minimum value (POS_min_) for a platform/phase. Incorporation of more complicated algorithms need to be supported by data availability and evidence from industry experts. POS_max_ and POS_min_ were defined in section b “Elicitation of probabilities of success”.

Robust evidence was not found about vaccines interdependence between platforms^5^.

5. “Operation Warp Speed” scaling up rule^6^.

The rule permits scaling up of “Operation Warp Speed” (OWS) logistics in case of insufficient portfolio R&D productivity and diversification outside of preselected lead candidates. The rule can be applied to a limited number of qualified candidates due to constrained resources. It will allow overlapping clinical trial phases, and speed up the development and manufacturing process, making “at risk” production for a broader range of development programs. The rule could be applied to selected Phase II candidates, including replacement of failing lead programs. Therefore, second round financing may not be significant as for the original cohort [25] assuming an already developed manufacturing and logistical ecosystem. Authors [8] limited the number of Phase III clinical trials to six. It seems contradictory to the industry practice where all Phase III trials are usually sped up.

Implementation of the proposed rule needs to be done gradually depending on portfolio risk profile, available resources, quality of candidates and policies.

### Modeling results

1. Analysis of baseline scenario.
  a. Vaccine approvals within 18 months.

Four scenarios were simulated – approvals within (1) 0-6 months; (2) 0-9 months; (3) 0-12 months, and (4) 0-18 months. Simulation results are presented in Figure 3.

**Figure 3.**
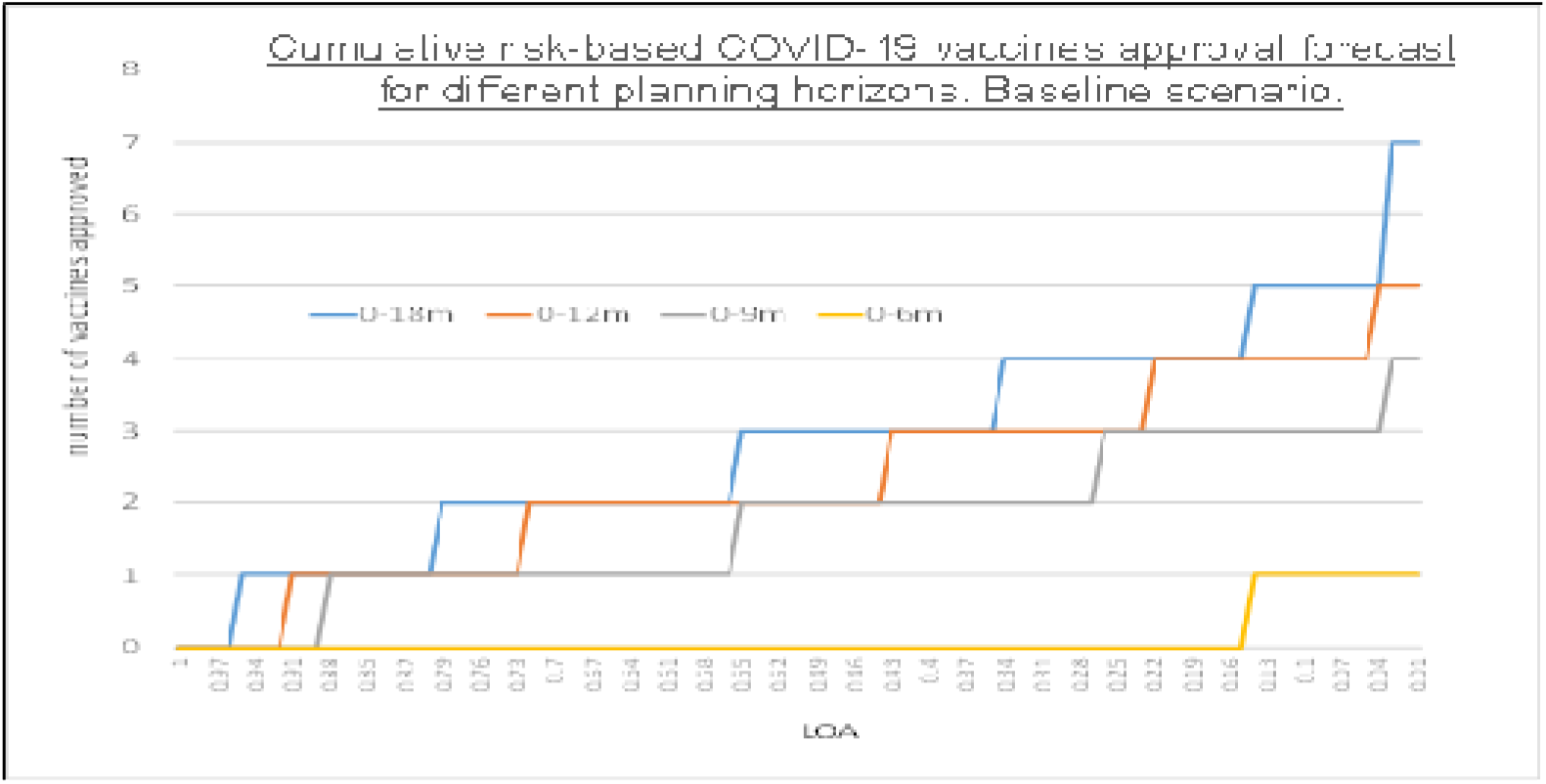
The graph shows maximum number of vaccine approvals (vertical axis) versus the likelihood of approvals (LOA) (horizontal axis). For example, in the 0 to 18-month period there is an almost 100% chance of at least one approval, approximately an 80% chance of more than two approvals, and approximately a 55% chance of more than three approvals (Shnaydman, 2020).

Modeling results indicate:

- For the planning period 0-6 months, LOA of one approval in low (about 15%);
- For the planning period 0-9 months, likelihood of zero approvals is about 10%, one approval is about 35%, two approvals is 30%, three approvals is about 21%, and four approvals is less than 5%. LOA for at least one approval is about 90%, LOA of two vaccines approval is 55%, etc.
- More favorable results are for planning periods 0-12 months and 0-18 months. There is a good chance that up to three-four vaccines will be approved in 0-18 month period.

In further calculations, planning period (0-18) months will be used to analyze portfolio productivity and risk for both R&D/clinical trials and manufacturing.

b. Allocation of vaccines approvals across technology platforms.

In the baseline scenario, the model forecasts that only vaccines from five platforms out of eight will be approved at the end of 2021 due to the longer duration of clinical trials and the chance of additional funding for remaining platforms. Number of approvals vs. LOA for each platform is presented on Figure 4. Platforms corresponding with approved vaccines are:

**Figure 4.**
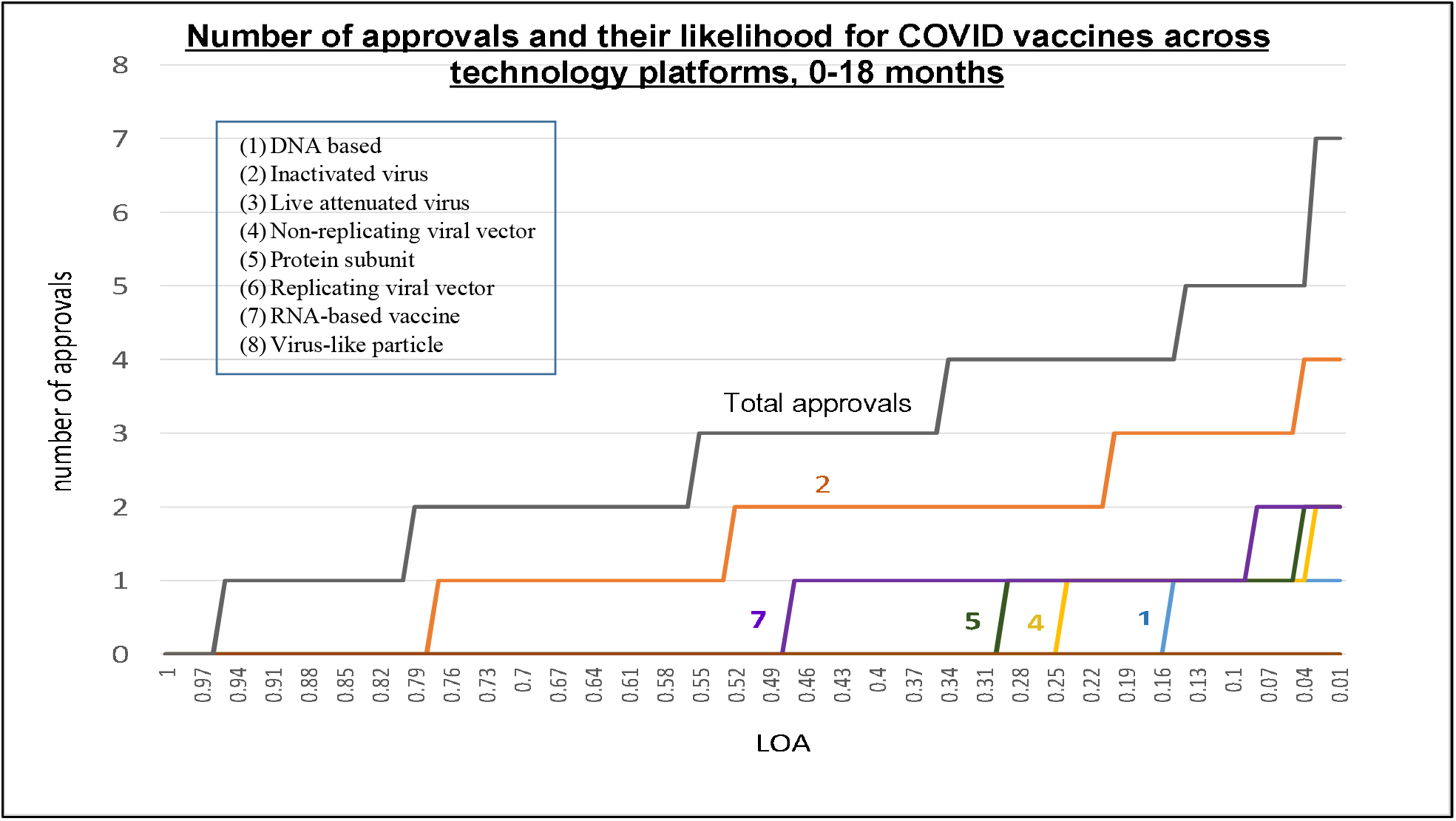
The graph shows maximum number of vaccine approvals (vertical axis) versus the cumulative LOA (horizontal axis) for the 0-18 month period across technology platforms. Results indicate that only vaccines from five platforms (1, 2, 4, 5, and 7) will be approved in 0-18 month period. Highest chances of approval has platform #2 (inactivated virus). Candidates from the WARP SPEED program [26] are among approved (Shnaydman, 2020).

- DNA based
- Inactivated virus
- Non-replicating viral vector
- Protein subunit
- RNA-based vaccine

Detailed analysis of each platform is presented in many sources, such as [8].

Simulation results presented in Figure 4 indicate that upgrades of manufacturing plants related to five platforms listed above should be prioritized. At the same time, according to [3], potential manufacturing capacity could be underutilized. How can utilization of available manufacturing capacity be maximized and production risk be minimized?^7^

c. Interdependence between vaccines within a platform.

Figure 5 analyzes sensitivity of the portfolio productivity to different interdependency rules.

**Figure 5.**
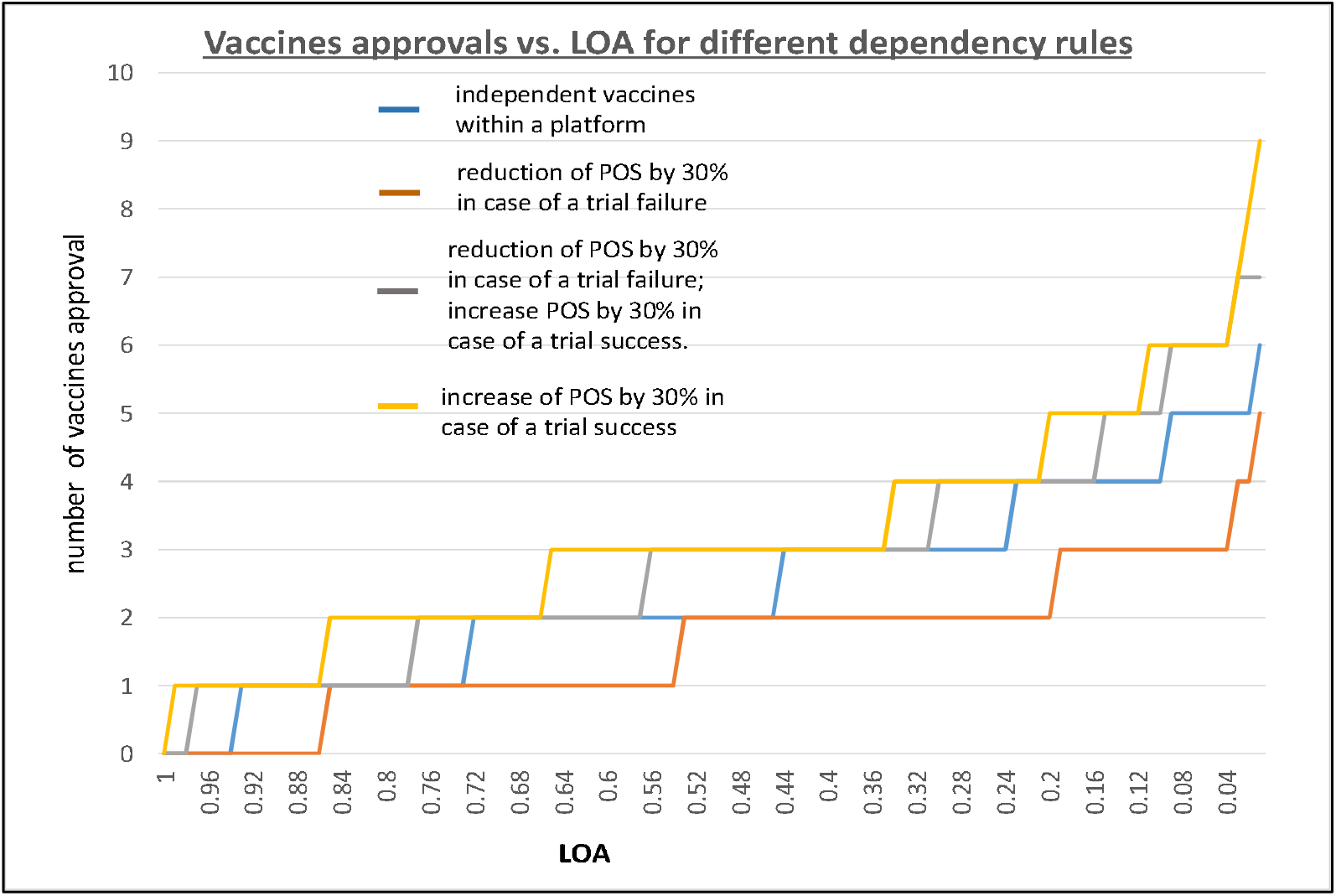
Sensitivity of portfolio productivity to interdependence rules. The graph shows number of vaccine approvals (vertical axis) versus the likelihood of approvals (LOA) (horizontal axis) for different degree of interdependence between vaccines within a platform. For example, reduction of POS by 30% in case of a trial failure, reduces portfolio productivity comparing to the baseline scenario (Shnaydman, 2020).

Figure 6 illustrates the POS dynamic for a phase/platform. POS is fluctuating depending on the outcome of previous trials within a platform.

**Figure 6.**
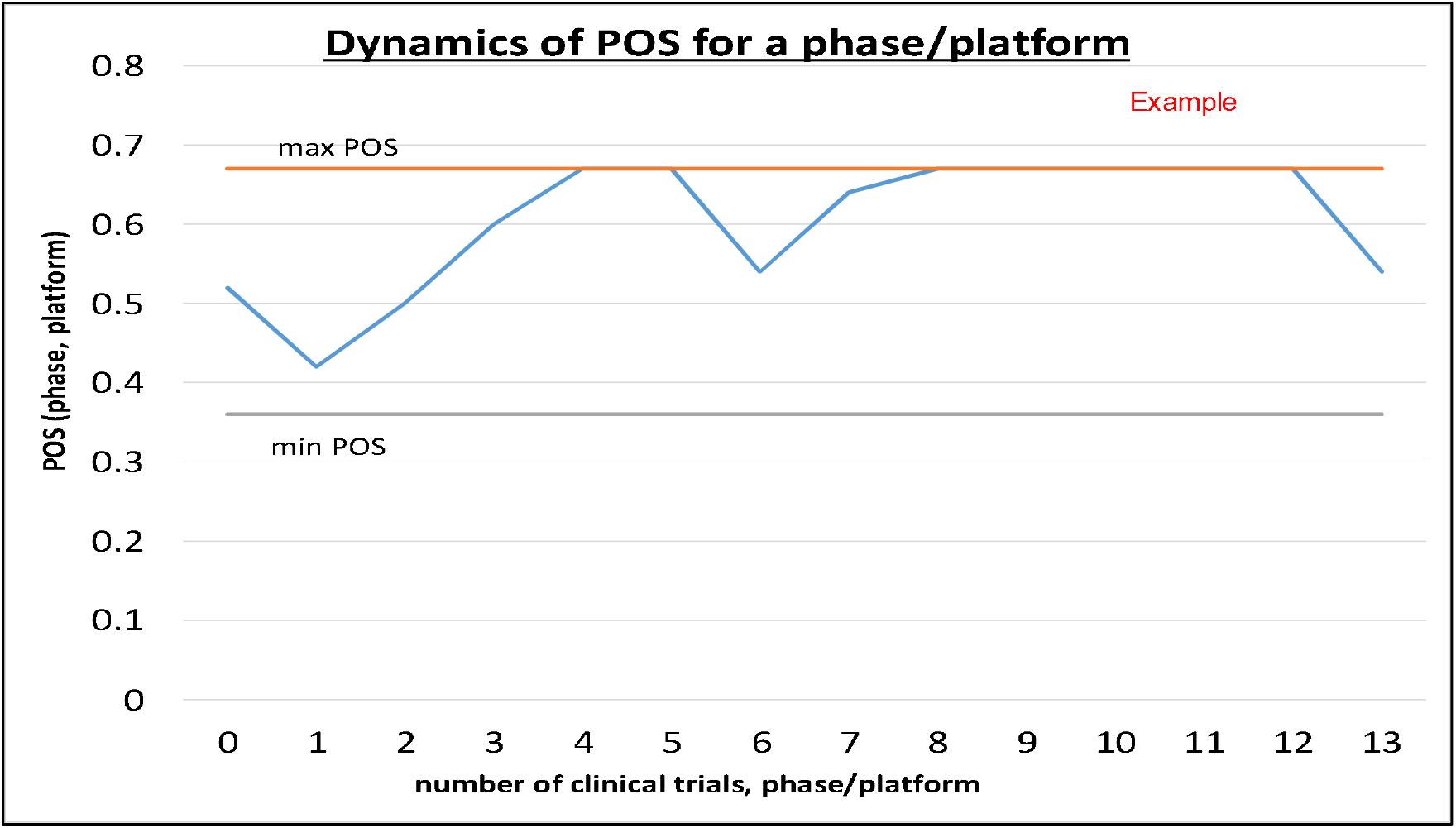
The graph shows fluctuations of POS (vertical axis) versus the number of clinical trials (horizontal axis) for a platform/phase due to vaccines dependency. In the presented scenario, first vaccine failed and POS reduced from 0.52 to 0.42. Then three successful trials raised POS to its maximum value 0.67 (Shnaydman, 2020).

d. R&D approval time and manufacturing time across platforms

Simulation results indicate that average completion of R&D cycle is around 5-8 months for five prioritized platforms. The R&D cycle time for vaccines in platforms “Live attenuated virus” (#3), “Replicating viral vector” (#6), and “Virus like particles” (#8) exceeds 18 months. Therefore, the manufacturing process will continue for about a year for five platforms to reach a short-term goal of two billion doses. Could short and long-term vaccine production goals be met? What would be the associated risk?

### Manufacturing

Vaccine manufacturers announced cumulative capacity that could produce up to one billion doses in 2020, and up to approximately nine billion in 2021 [3, 25], but in reality, due to high attrition rates, some candidates may fail and corresponding capacity plans may be altered. Therefore, the model will address several questions, such as:

- Would R&D productivity of vaccines pipeline be sufficient to meet world-wide vaccination demand in the shortest time period?
- Would manufacturing capacity be sufficient to produce enough vaccine doses worldwide?
- Would corresponding production risk of not meeting the goals be tolerable?

*Production risk* is defined as probability of not meeting vaccine demand. The higher demand, the higher the risk of meeting the goal for both end of 2021 (two billion doses) and vaccination of the world (11.2 billion doses).

#### Short-term goal – two billion doses produced by the end of 2021

Figure 7 indicates that the risk of not meeting short-term production goal of two billion doses is relatively high (about 50%). It will be much higher for the world vaccination. How can the production risk be mitigated and the portfolio risk profile be improved in order to meet vaccination goals?

**Figure 7.**
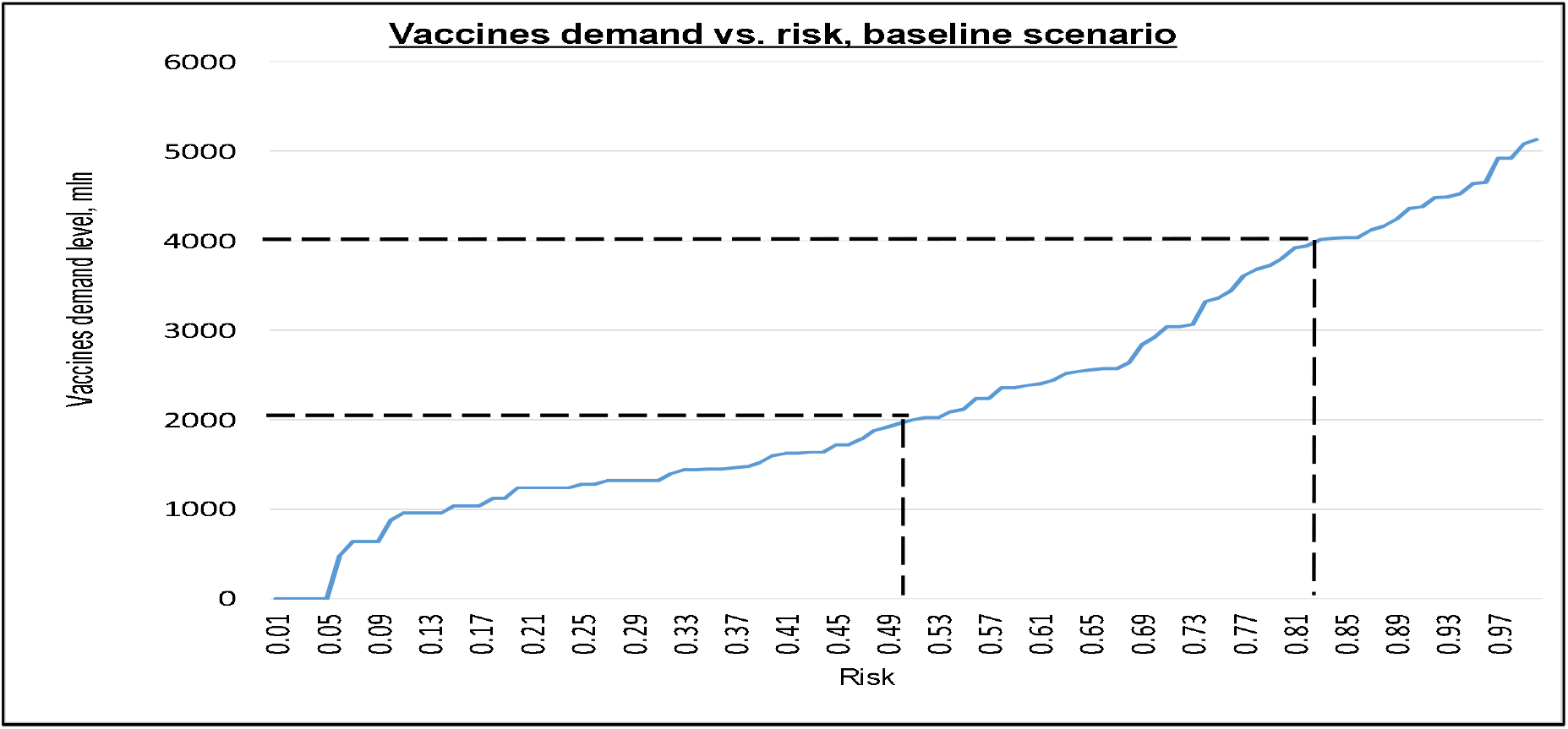
The graph shows how risky manufacturing production is for different goals – two billion and four billion doses within the 0-18 month time interval. There is approximately a 50% chance that the goal of two billion doses will be met, and only 18% probability, approximately, for four billion doses (82% risk). X axis is risk, Y axis is the vaccine demand level, in millions of doses (Shnaydman, 2020).

Figure 8 presents the dynamics of average vaccine production.

**Figure 8.**
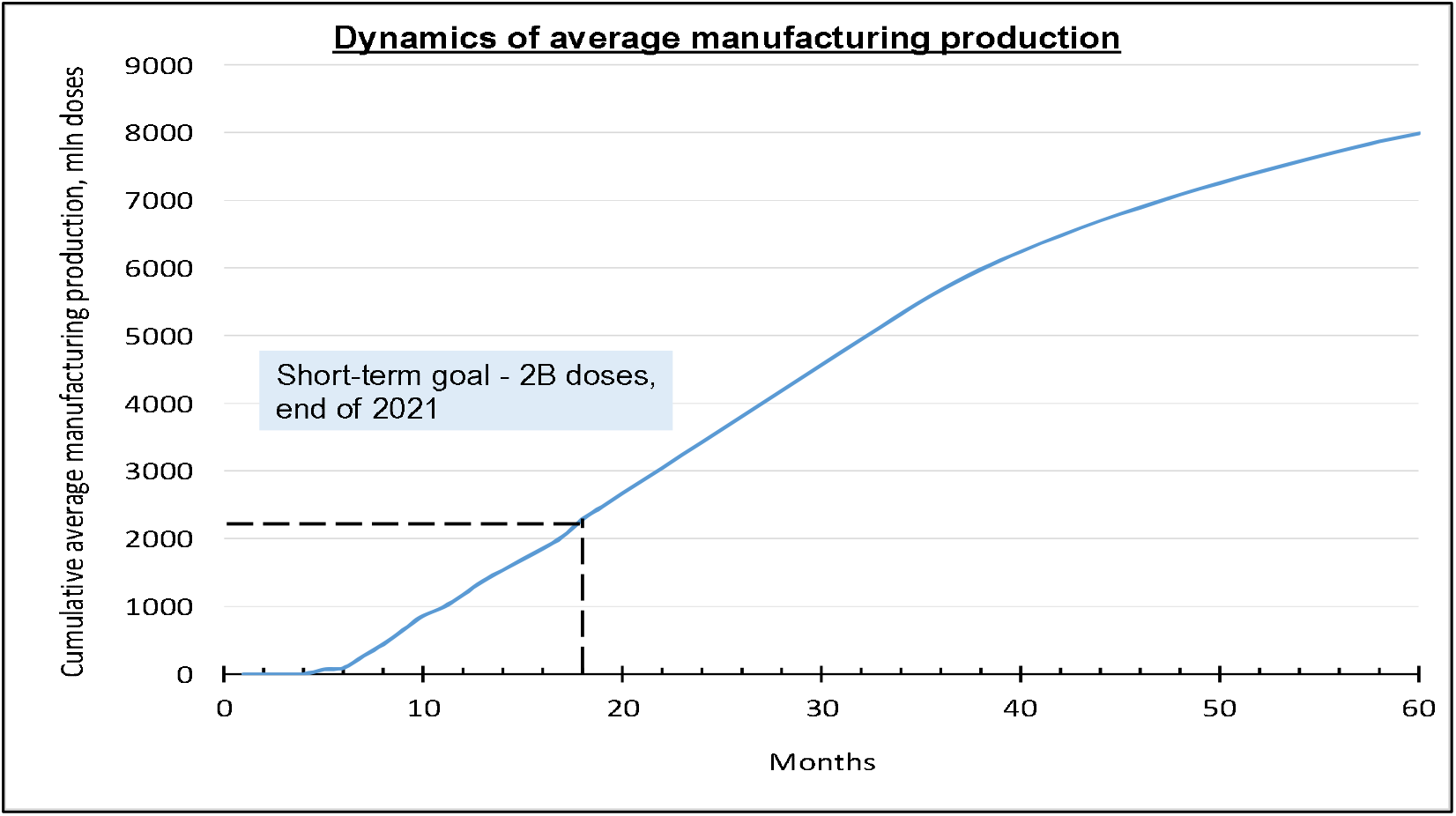
The graph shows average cumulative production dynamics for COVID vaccines, 0-18 months. On average 2.2 billion doses could be produced at the end of 2021. X axis is time in months, Y axis is the cumulative vaccine production, in millions of doses (Shnaydman, 2020).

Average dynamic production of 2.2 billion doses means that the risk of meeting the short-term goal is about 50%. How can this risk be reduced?

### Risk mitigation

Several potential portfolio mitigation strategies were analyzed in [4]. They included: (1) Increased portfolio size; (2) Compressed cycle times; (3) POS increase by 10%. This paper will expand the number of risk mitigation strategies to guarantee world vaccination in the shortest time and with minimum risk.

#### 1. Increasing manufacturing productivity

Simulation results presented on Figure 9 indicate that an increase in manufacturing capacity corresponds with only a marginal increase in vaccine production.

**Figure 9.**
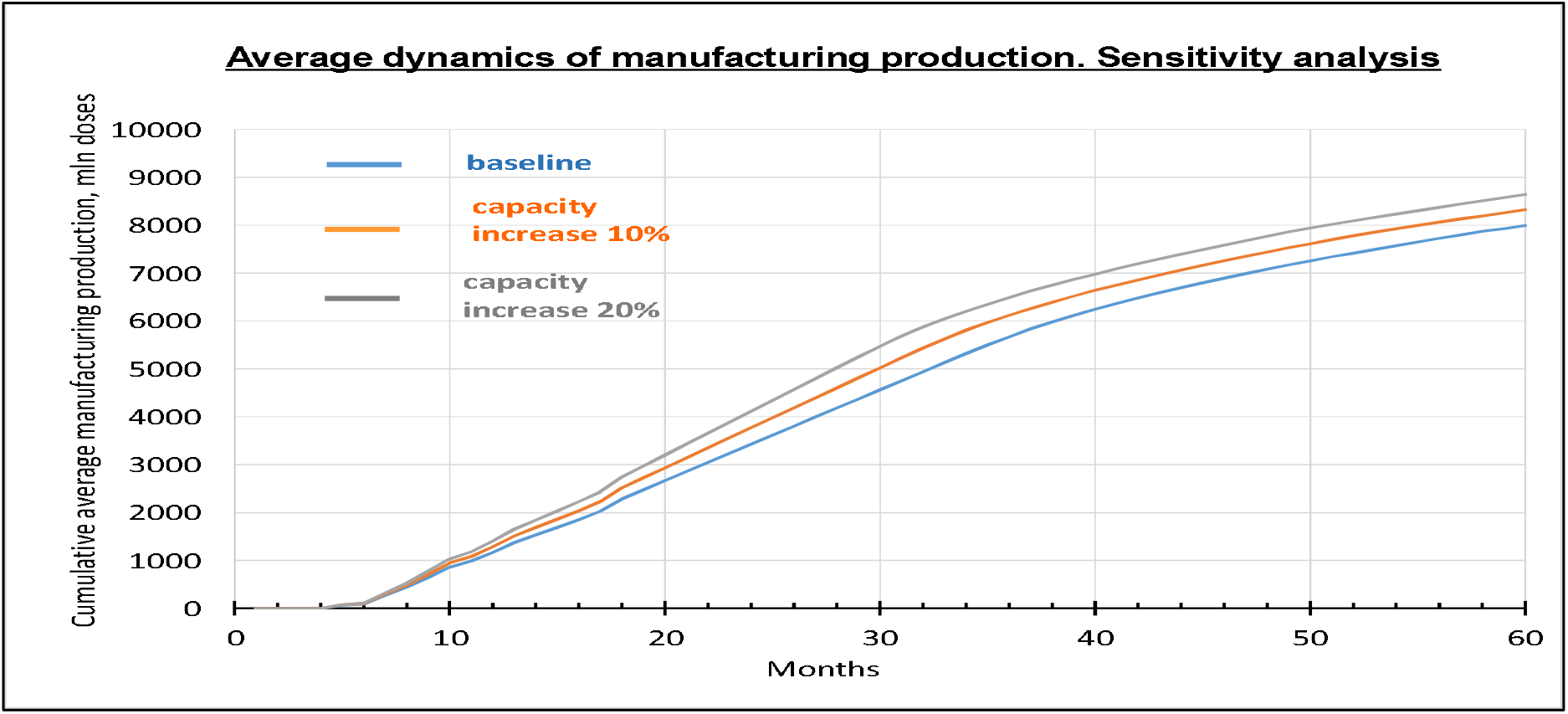
The graph shows that increasing manufacturing productivity by up to 20% does not significantly increase production. For example, at the end of 2021, cumulative production will increase from 2.2 to 2.5 billion doses, if manufacturing capacity increases by 20%. X axis is time in months, Y axis is cumulative vaccine production, in millions of doses (Shnaydman, 2020).

Therefore, it appears that vaccine manufacturing productivity is not a system bottleneck. Is there a more effective solution? Deeper analysis indicates that R&D portfolio productivity could be a bottleneck. How can R&D productivity be increased?

#### 2. Scaling of OWS

Companies benefiting from the OWS^8^ compressed and overlap clinical trials cycle times are based on significant US government funding. These companies became leaders in the vaccine race, and, if their vaccines are successful, they may produce enough doses to meet worldwide demand. At the same time, the risk of not meeting production goals is still significant (see Figure 7).

How can the vaccine portfolio production risk profile be improved?

In this paper, the strategy based on scaling of OWS was simulated.

According to Figure 10, number of vaccines approvals significantly increases comparing to the baseline scenario. Portfolio risk profile is also more favorable than in the baseline scenario.

**Figure 10.**
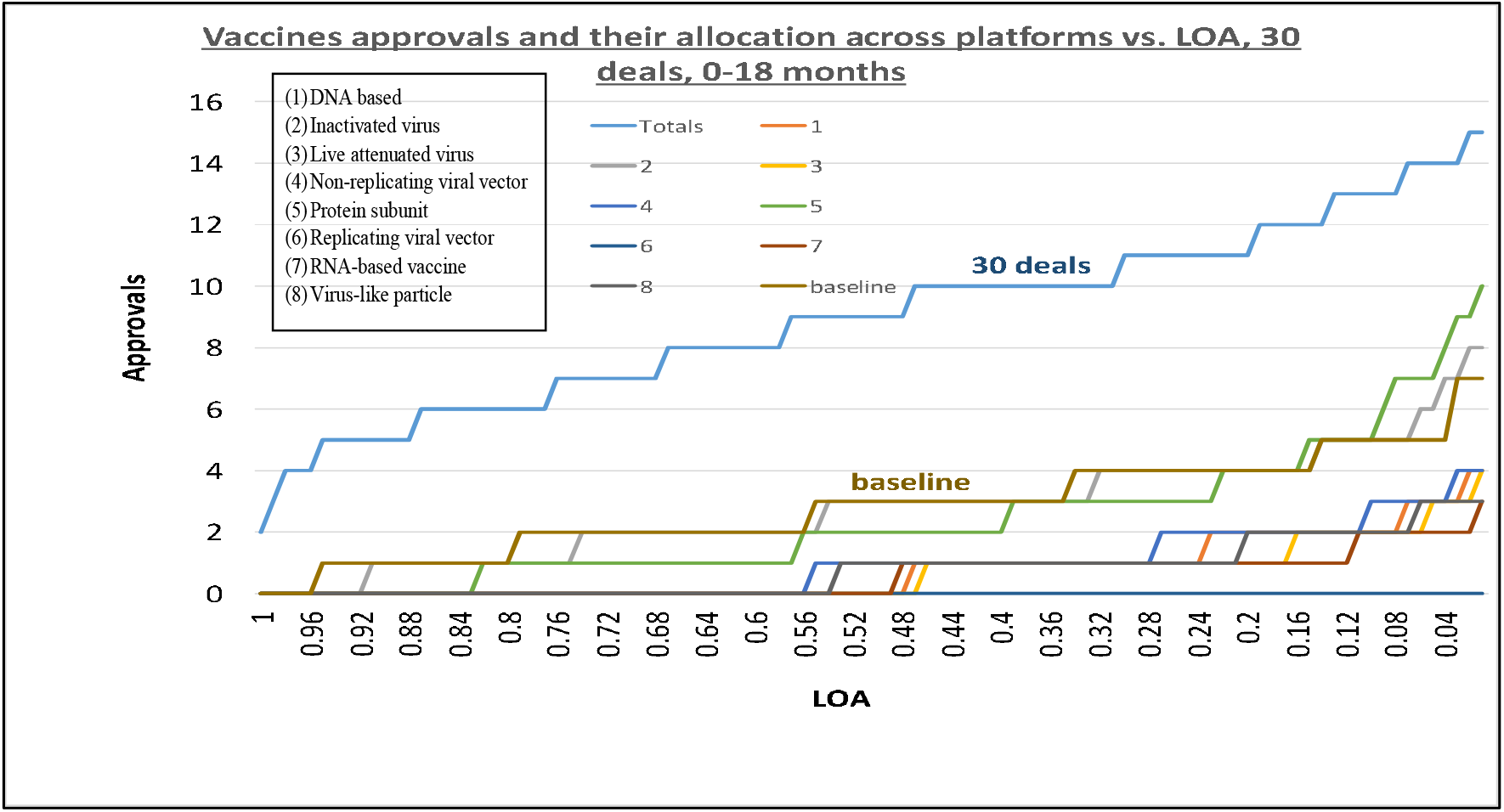
The graph shows that 30 collaborative agreements/sponsorships based on governmental and international funding significantly increase LOA of approvals comparing with baseline scenario. Median approvals for collaborative scenario = 9 versus a baseline scenario = 3. Also, vaccines from seven platforms are contributed in a “collaborative” scenario versus vaccines from five platforms in a baseline scenario, due to compression of clinical trials cycle times and at risk execution of Phase III clinical trials. X axis is LOA, Y axis is number of approvals (Shnaydman, 2020).

Worldwide vaccines production risk profiles for different risk mitigation scenarios are presented in Figure 11.

**Figure 11.**
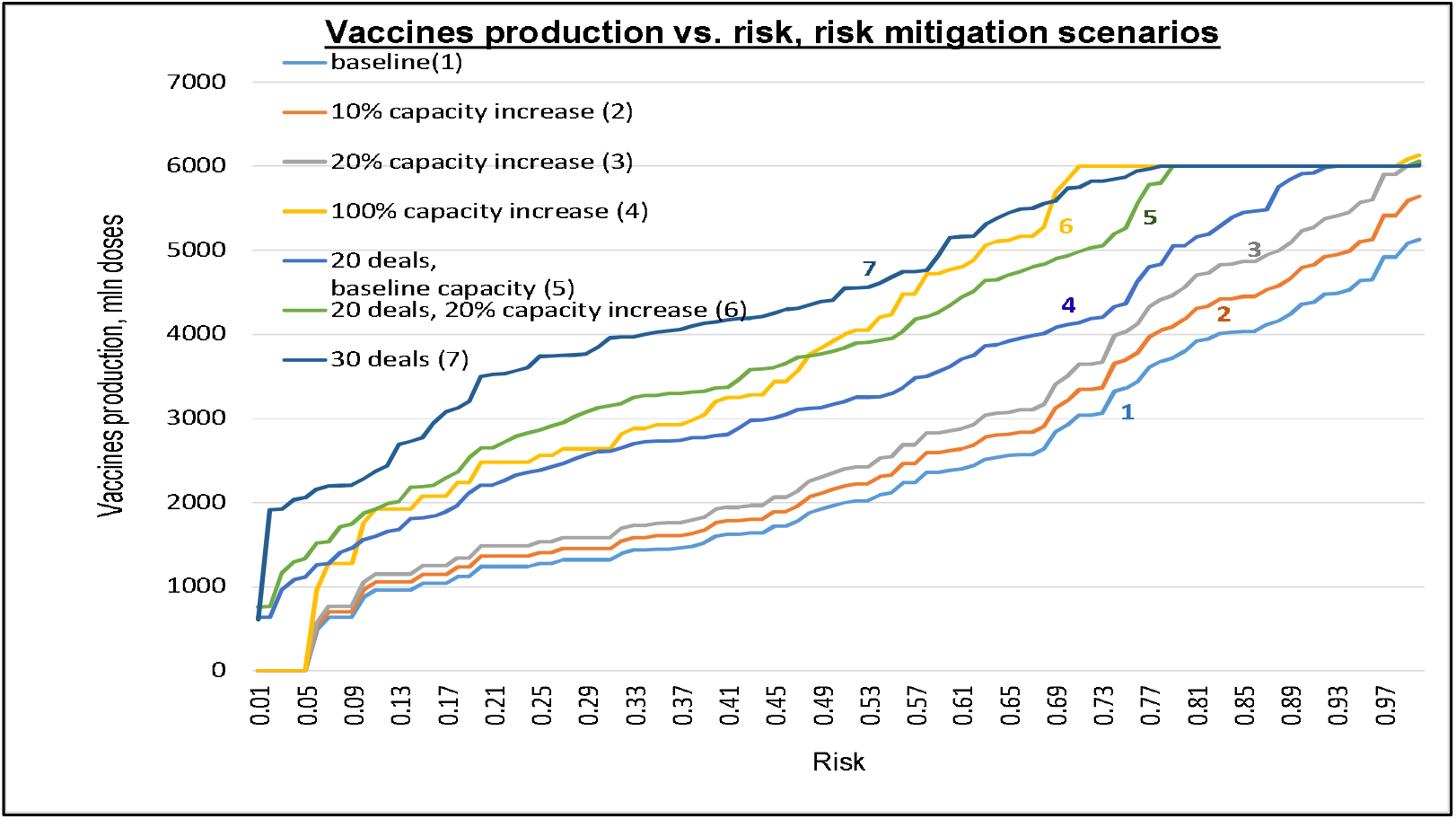
The graph shows that mitigation strategy corresponding to increasing number of deals (for example, R&D subsidizing, buy-outs, in-licensing) can significantly reduce production risk when compare to just mitigation strategy related to increasing manufacturing capacity only. X axis is production risk, Y axis is cumulative vaccine production, in millions of doses. Figure 11 indicates that in “collaborative” scenario with 30 deals (for 200+ portfolio candidates) the risk of reaching two billion doses is close to zero; the risk will be reduced from 90% to about 35% for the goal of producing four billion doses (Shnaydman, 2020).

Figure 12 shows the risk adjusted (average) dynamics of the production of multiple vaccines.

**Figure 12.**
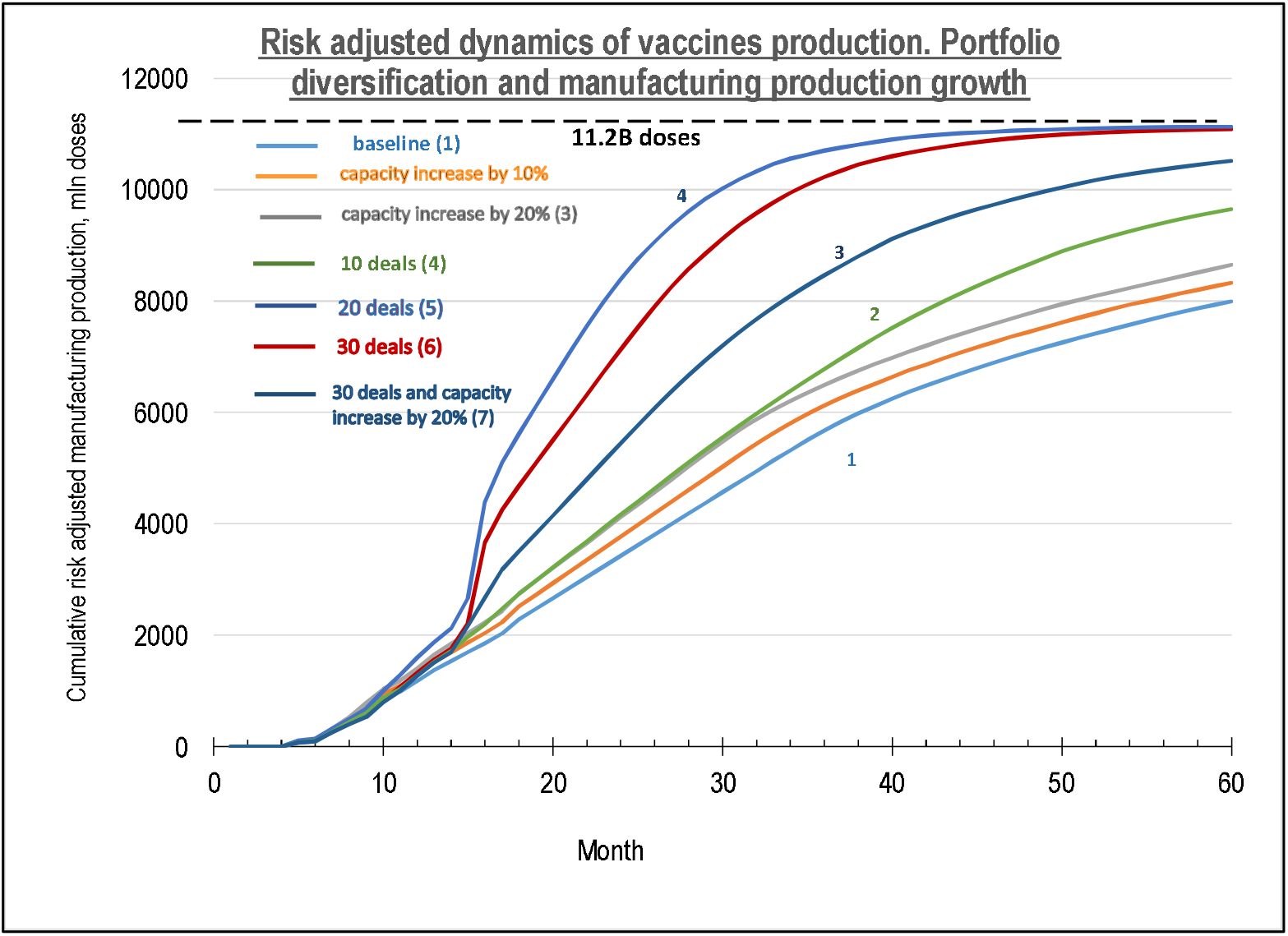
The graph shows that partnering deals increase vaccines average production volume from 2.4B to 4.5B by the end of 2021 if “partnering” mitigation strategy would be implemented (month 18) (Shnaydman, 2020).

Production variation increases between months 20 and 40 among the scenarios due to delayed Phase III entrance.

The likelihood of worldwide vaccination in 36 months is high as shown in Figure 13 if global policy is focused on portfolio diversification and financing of the most promising candidates beyond original OWS.

**Figure 13.**
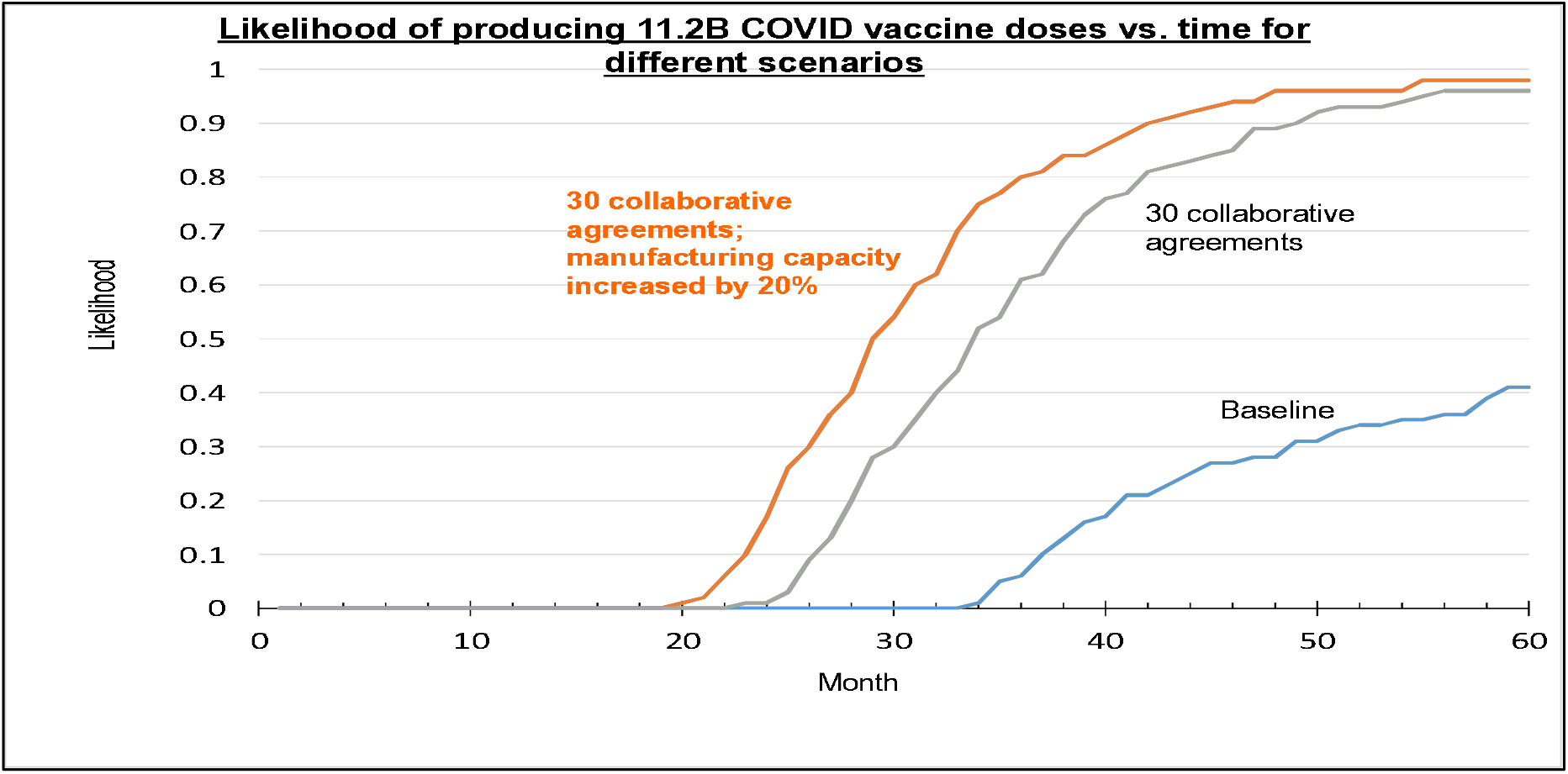
The graph shows that prioritization of vaccine candidates for scaled up OWS can significantly increase likelihood of worldwide vaccination. World vaccination could be accomplished in 36 months with low risk (in case of smooth distribution). For the baseline scenario, likelihood of worldwide vaccination even in 60 months could be only 40% (Shnaydman, 2020).

### Assessment of economic impact of proposed portfolio diversification strategy

In order to prove effectiveness of proposed COVID vaccines portfolio diversification strategy, the timing of the production of 11.2 billion doses was estimated by crude extrapolation of curves (1-4) in Figure 12.

In order to do that let’s make several assumptions:

1. World GDP = US$ 87.7 trillion in 2019 [29].
2. Historical world GDP annual growth is about 3.5%^9^ [30].
3. World vaccination will start almost immediately after approvals of vaccines (beginning of 2021). Some vaccination delays are possible, but not significant.
4. World economic growth will be on track if about 70% of the world population will be vaccinated.
5. Four scenarios were analyzed. They are marked on Figure 12 as 1 – baseline; 2 – 10 deals; 3 – 20 deals and 4 - 30 deals and increased manufacturing capacity by 20% (most favorable scenario).

Figure 14 illustrates assessment technique.

**Figure 14.**
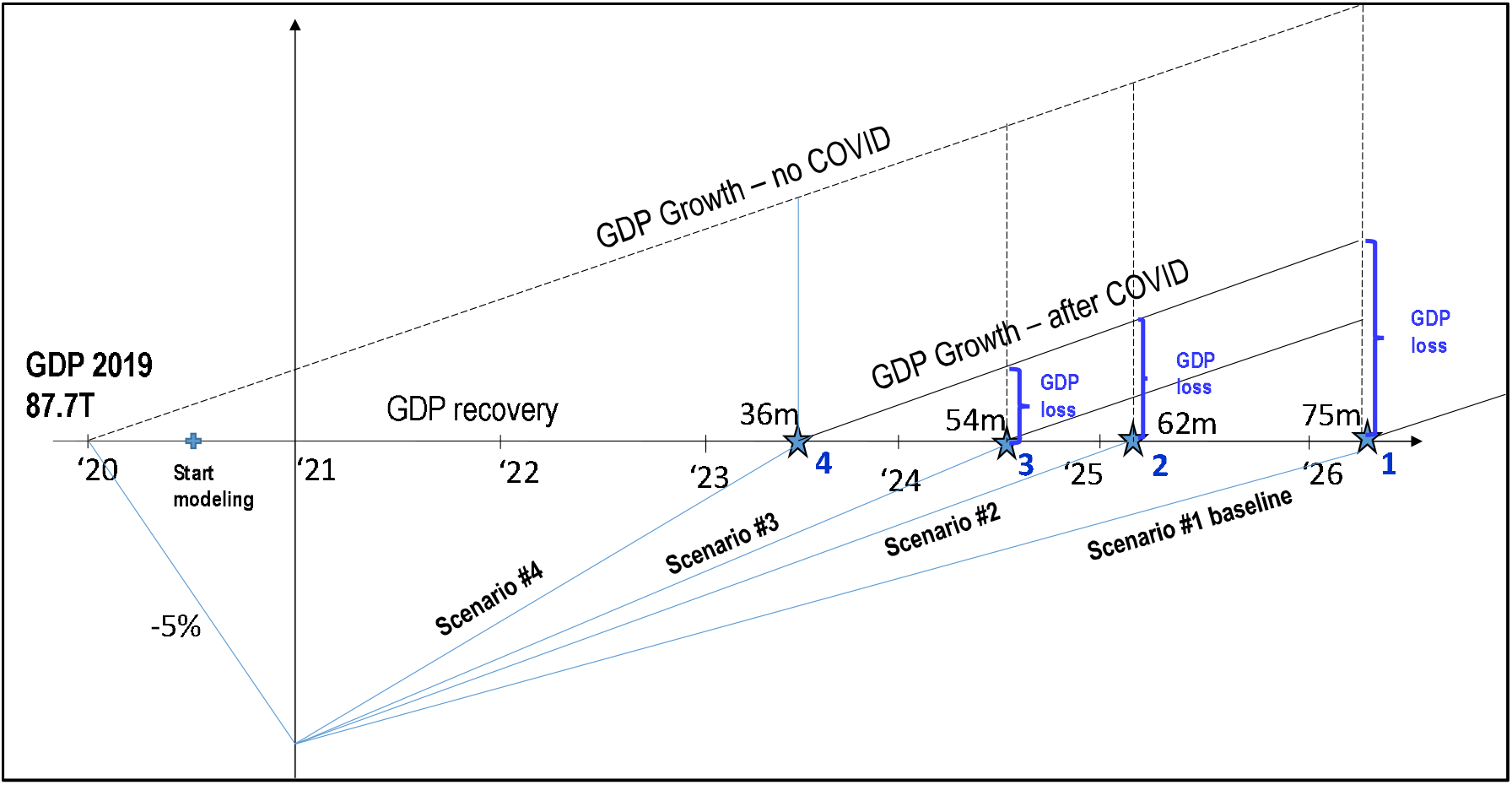
The diagram illustrates calculation of the World GDP loss for less favorable scenarios than Scenario #4. In 2020, world GDP contracted by about 5% [. In the best scenario, it is assumed that world economy will come back to pre-COVID-19 state in 36 months (world vaccination), and GDP loss=0 comparing to the baseline scenario – 75 months (extrapolated) where GDP loss would be maximal (Shnaydman, 2020).

According to the assessment, in order to minimize GDP loss, additional investments should be made to speed up Phase III trials and maximize portfolio productivity. The graph on Figure 15 shows the potential impact of global vaccine development strategies on lost world GDP.

**Figure 15.**
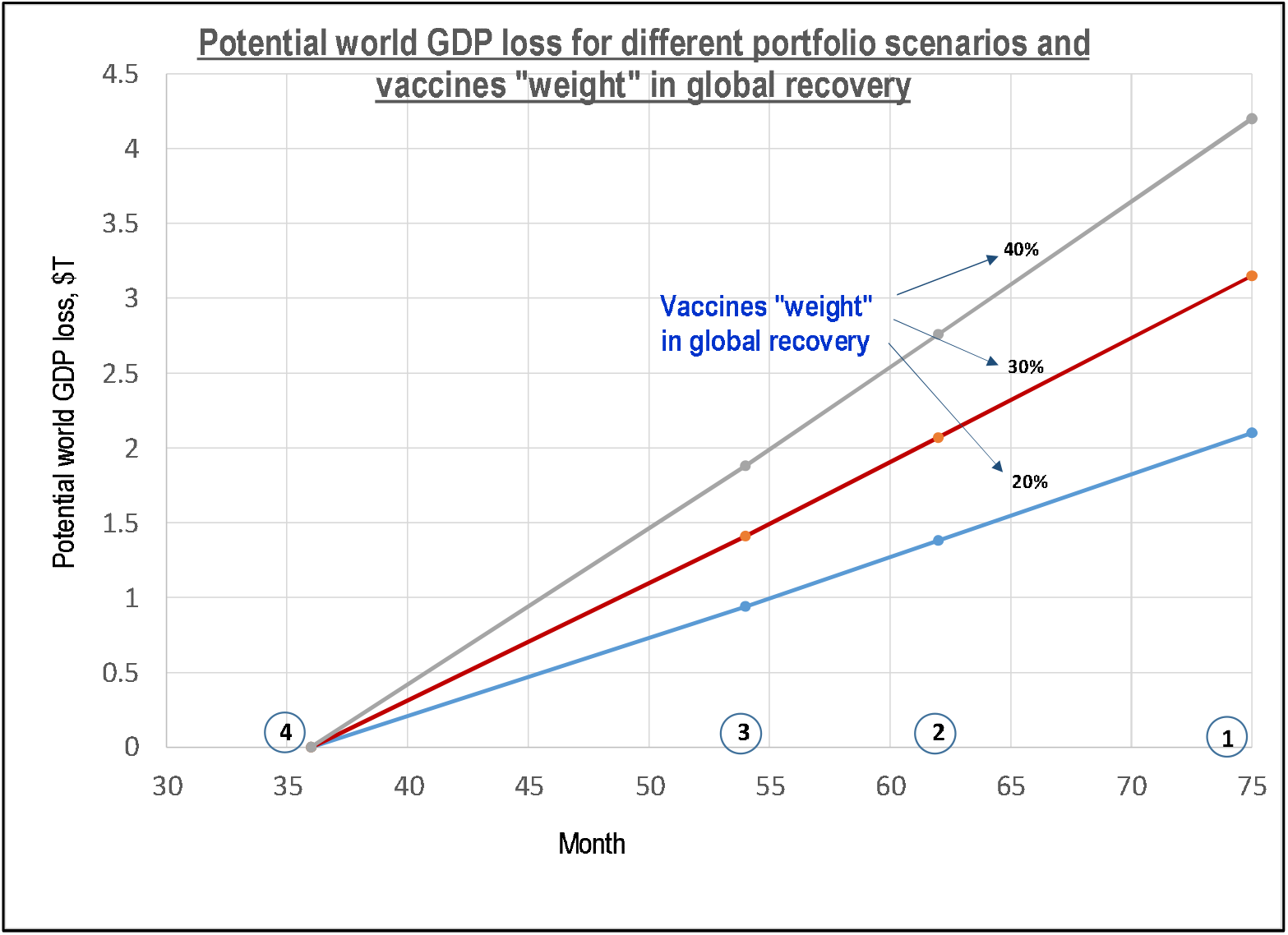
The graph shows impact of vaccine development strategies on potential world GDP loss for different vaccines’ “weight” in world recovery. For example, even if the world vaccination will contribute 20% to the GDP recovery, GDP loss for Scenarios 1-3 will range from ∼0.9T to 2.1T comparing to the “best case” scenario. For 40% world vaccination weight global GDP loss could reach $4.2T (if the “best” scenario is not materialized), for US it could be about $1.05T over three years. X axis – time in months, Y axes – Potential world GDP loss (Shnaydman, 2020).

ACT Accelerator [31] estimates that $35 billion is required to meet demand in all COVID-19 related products. If several billion dollars will be invested in OWS scale up, return on investment would be significant.

## Supporting information

Input data file

## Data Availability

Data is available

## Acknowledgement

The author is very thankful to Michael Quinn.

## Conclusion

The paper presents a methodology and modeling results for COVID-19 vaccines portfolio forecasting, including R&D output (rate of approvals and their likelihood at a platform level) and manufacturing production output. Methods of analysis include Monte-Carlo portfolio simulation and a technique for elicitation of probabilities of success (POS) at a platform level. Data was extracted from multiple sources.

Simulation results for a baseline scenario indicate that time to produce enough vaccines for world vaccination is significant despite prioritization of a handful portfolio candidates as a part of Operation Warp Speed (OWS). Limited manufacturing capacity and insufficient portfolio diversification contribute to that.

In order to minimize the time and risk of world vaccination, scaling up of OWS could be very beneficial including additional financing in both Phase III clinical trials and manufacturing for additional vaccines development programs. It would lead to a reduction in global production time for world vaccination from 75 months for baseline scenario to 36 months, reducing potential global GDP loss up to $4.2 trillion (US - $1 trillion) when compared to the baseline scenario.

1 More accurate numbers will be probably available within several months.

2 Input data presented in the Appendix – Model_data_102020.xls

3 Some LOA benchmarks for COVID-19 platforms were presented in [3].

4 Typically, x and y are in range 20-30% based on the industry practice.

5 Further research may be needed.

6 The program could be international

7 Please, see “Risk mitigation” section for one of possible solutions.

8 And other programs

9 Calculated for years 1961-2019.

